# ApoA-1 versus HDL-C as Markers of Cardiovascular Risk

**DOI:** 10.1101/2025.09.09.25335464

**Authors:** Selin Bilgic, Karol Pencina, Michael Pencina, Line Dufresne, George Thanassoulis, Allan Sniderman

## Abstract

**Background:** Conflicting results have been reported as to the relative importance of apoA-1 versus HDL-C as markers of ASCVD risk.

**Methods:** Residual discordance analysis with Cox proportional hazard models comparing apoA-1 and HDL-C as markers of ASCVD risk was applied to a sample of 291,995 UK Biobank, followed for a median of 11 years. Interaction test for the two markers and estimation of the effects of partitioning HDL-C into apoA-I, log-triglyceride and the remaining residual were also performed.

**Results:** ApoA-1 and HDL-C had similar associations with ASCVD risk (HRs of 0.85, p-value < 0.001 for both). The residual of HDL-C added significantly to the risk associated with apoA-1 as did the residual of apoA-1 to HDL-C. There was a statistically significant interaction between apoA-I and HDL-C (HR**=**1.05, 95% CI: (1.04, 1.06); p < 0.001). Decomposing HDL-C into the 3 components, apoA-I accounted for the largest portion of the effect with a HR of 0.85 95%CI: (0.83, 0.86) with smaller effects for lnTG: 1.04 (1.02, 1.06) and residual of HDL-C: 0.98 95%CI: (0.96, 0.995).

**Conclusions:** HDL-C and apoA-1 have associations of equivalent strength with ASCVD risk with significant interaction modifying the effect of one by the other. Upon decomposition, ApoA-I retained more of the effect of HDL-C as compared to log-triglycerides. While only observational, the results are consistent with the relation of HDL to risk not being determined by the concurrent level of triglyceride.

**Clinical Perspective:** The plasma levels of HDL-C and apoA-I are potent predictors of cardiovascular risk. However, there are few data comparing the relative precision of HDL-C and apoA-I for this purpose. Moreover, the risk of low HDL-C has been attributed to concurrent hypertriglyceridemia, consequently downgrading the potential importance of HDL in predicting or explaining risk.

**Novel Findings:** Based on residual discordance analysis, HDL-C and apoA-I have similar predictive precision for ASCVD risk. However, each adds significantly to the other and. Moreover, triglycerides account for only a small portion of the risk attributable to HDL with apoA-I accounting for the principal portion.

**Clinical Significance:** HDL, whether measured as HDL-C or apoA-I, is a potent predictor of ASCVD risk. It remains essential to search for the biological basis or bases for these relationships.

## Introduction

Both the apolipoprotein B (apoB) and apolipoprotein A (apoA) lipoproteins are powerful predictors of cardiovascular risk. ^1–3^ Cardiovascular risk increases as the concentrations of LDL-C, non-HDL-C, or apoB in plasma increase but decreases as the concentrations of HDL-C or apoA-1 in plasma increase. While a causal role in atherogenesis has been firmly established for the apoB lipoproteins, no similar strength of evidence has been gathered for the apoA lipoproteins. ^4,5^ On the contrary, multiple Mendelian randomization analyses have concluded no causal role exists for HDL-C ^6–8^ and no clinical benefit has been observed from pharmacological therapies, which elevate HDL-C. ^9^ Notwithstanding this array of evidence against causality, the fact remains that the concentration of HDL in plasma, whether measured as HDL-C or apoA-I, predicts cardiovascular risk consistently and powerfully.

However, the issue is complicated because of the inverse relation between plasma triglycerides and HDL-C due to core lipid exchanges between VLDL and LDL particles mediated by cholesterol ester transfer protein. Because the evidence for a causal role in atherogenesis is stronger for VLDL than for HDL, it has been argued that this drives the inverse relation of HDL-C to risk and, accordingly, it is unnecessary to adjust for HDL in determining the risk attributable to VLDL. Beyond this, results have differed as to which of the two principal components of HDL–the mass of cholesterol, HDL-C or the concentration of the major apolipoprotein in HDL particles, apoA-I–is the more precise index of the cardiovascular risk.^10,11^

Therefore, the objectives of the present study are: first, to compare HDL-C and apoA-1 as markers of cardiovascular risk and then, to estimate the role of plasma triglycerides as determinants of the inverse relations of HDL-C and apoA-I to risk. Because HDL-C and apoA-I are highly correlated variables, residual discordance analysis will be the primary analytical tool to examine their respective relationships to cardiovascular risk.

## Methods

The initial cohort of 502,413 participants in UK Biobank was filtered to exclude individuals with CVD or who were taking lipid-lowering therapy at baseline examination. Additionally, those with missing records for HDL-C, apoA-I, LDL-C, non-HDL-C, apoB, triglycerides, HbA1c, BMI, SBP, sex, smoking history, hypertension treatment, or diabetes treatment were excluded. Participants with triglycerides ≥ 400 mg/dL, apoB <20 mg/dL and > 400 mg/dL, or LDL-C ≥ 250 mg/dL were also excluded, resulting in a final analytic sample of 291,995 participants.

All statistical analyses in this cohort were performed using R version 4.2.2. Residuals of HDL-C and apoA-I were created to represent the amount of HDL-C that is not explained by apoA-I and the amount of apoA-I that is not explained by HDL-C. Regressing HDL-C on apoA-I formed the HDL-C residual; regressing apoA-I on HDL-C created the apoA-I residual. The distribution of triglycerides was skewed in the sample; therefore, the triglyceride variable was natural log-transformed (ln TG). Pearson correlation coefficients were calculated between HDL-C, LDL-C, non-HDL-C, ln TG, apoB, apoA-I, the HDL-C residual, the apoA-I residual and the HDL-C/apoA-I ratio. Next, two main Cox proportional hazards models were created to estimate the effects of HDL-C, apoA-I and their interaction on ASCVD incidence. Due to the high correlation between HDL-C, apoA-I, we modelled separately the effects of HDL-C, or the apoA-I residual while the other included apoA-I and the HDL-C residual. Two additional Cox proportional hazards models were created to better understand the relationship between HDL-C, TG and apoA1. The first one was a fully adjusted model with HDL-C while the other was a fully adjusted model which partitioned HDL-C into apoA1, lnTG and the residual of HDL-C when regressing it on apoA1 and lnTG. All models were adjusted for SBP, HbA1c, BMI, age, sex, hypertension treatment, diabetes treatment, and smoking status. Hazard ratios (HR), which were expressed per 1 standard deviation, 95% confidence intervals (CI), and p-values were produced.

## Results

### A. General Findings

In the main cohort of 291,995 participants, the median age at baseline was 56 years, 42.1% of participants were male, and there were 19,849 ASCVD events over a median follow-up of 11 years (Table 1). The median values for HDL-C, apoA-I, non-HDL-C, LDL-C, and apoB were 55.7 mg/dL, 153.0 mg/dL, 167.5 mg/dL, 141.6 mg/dL, and 104.8 mg/dL, respectively (Table 1).

**Table 1.** Participant Characteristics of Main UK Biobank Cohort.

| Variables | Values/Median [SD] (%) |
| --- | --- |
| **N** | 291995 |
| **Age (years)** | 56.00 [8.11] |
| **Male (N)** | 123057 (42.1) |
| **Incidence ASCVD (N)** | 19849 (6.8) |
| **Incidence ASCVD in Males (N)** | 11678 (4.0) |
| **Incidence ASCVD in Females (N)** | 8171 (2.8) |
| **Follow-up (years)** | 10.96 [2.06] |
| **Body Mass Index (kg/m<sup>2</sup>)** | 26.27 [4.59] |
| **Systolic Blood Pressure (mmHg)** | 135.00 [18.61] |
| **Blood Pressure Medication (N)** | 36020 (12.3) |
| **Ever Smoked (N)** | 170628 (58.4) |
| **HbA1c (%)** | 5.33 [0.46] |
| **Diabetes Medication (N)** | 2379 (0.8) |
| **Total Cholesterol (mg/dL)** | 225.06 [39.73] |
| **HDL-C (mg/dL)** | 55.68 [14.19] |
| **HDL-C residual** | -0.05 [1.00] |
| **Non-HDL-C (mg/dL)** | 167.52 [38.18] |
| **LDL-C (mg/dL)** | 141.57 [30.66] |
| **TG (mg/dL)** | 125.59 [71.17] |
| **ApoB (mg/dL)** | 104.80 [22.67] |
| **ApoA-I (mg/dL)** | 153.00 [26.89] |
| **ApoA-I residual** | -0.04 [1.00] |
| **HDL-C:apoA-I Ratio** | 0.36 [0.04] |

The Pearson correlation coefficient between HDL-C and apoA1 was 0.92 (Table 2). Thus, about 85% of one variable can be explained by the other, leaving 15% unexplained. By contrast, there were no clinically significant correlations between either apoA-I or HDL-C and apoB, LDL-C or non-HDL-C. As expected, there was a moderately strong inverse relation between HDL-C and lnTG (r=0.47), a relation that was considerably stronger than between apoA-I and lnTG (r=0.28). Similarly, although the residuals of apoA-I and HDL-C were highly correlated, the relations to lnTG were stronger for the HDL-C residual (r=0.54) than for the apoA-I residual (r=0.39). As anticipated, the HDL-C/apoA-1 ratio was inversely and moderately related to plasma triglycerides The HDL-C/apoA-I ratio was also moderately related to the level of apoA-1 (r=0.51) but more strongly related to the level of HDL-C (0.80).

**Table 2.** Pearson Correlation Coefficients in the Main UK Biobank Cohort.

|  | HDL-C | Non-HDL-C | ApoB | LDL-C | lnTG | HDL-C/ApoA-I | ApoA-1 Residual | HDL-C Residual |
| --- | --- | --- | --- | --- | --- | --- | --- | --- |
| ApoA-1 | 0.92<br>($p<0.001$) | -0.02<br>($p<0.001$)<br>) | -0.08<br>($p<0.001$)<br>) | 0.04<br>($p<0.001$) | -0.28<br>($p<0.001$) | 0.51<br>($p<0.001$) | 0.39<br>($p<0.001$) | 0<br>($p=1.00$) |
| HDL-C | | -0.07<br>($p<0.001$)<br>) | -0.11<br>($p<0.001$)<br>) | 0.02<br>($p<0.001$) | -0.47<br>($p<0.001$) | 0.80<br>($p<0.001$) | 0<br>($p<0.001$) | 0.39<br>($p<0.001$) |
| Non-HDL-C | | | 0.96<br>($p<0.001$)<br>) | 0.98<br>($p<0.001$) | 0.51<br>($p<0.001$) | -0.12<br>($p<0.001$) | 0.12<br>($p<0.001$) | -0.14<br>($p<0.001$) |
| ApoB | | | | 0.96<br>($p<0.001$) | 0.42<br>($p<0.001$) | -0.11<br>($p<0.001$) | 0.06<br>($p<0.001$) | -0.10<br>($p<0.001$) |
| LDL-C | | | | | 0.38<br>($p<0.001$) | -0.01<br>($p<0.001$) | 0.07<br>($p<0.001$) | -0.05<br>($p<0.001$) |
| lnTG | | | | | | -0.62<br>($p<0.001$) | 0.39<br>($p<0.001$) | -0.54<br>($p<0.001$) |
| HDL-C/<br>ApoA-I |  |  |  |  |  |  | -0.58<br>(p<0.001) | 0.84<br>(p<0.001) |
| ApoA-I<br>Residual |  |  |  |  |  |  |  | -0.92<br>(p<0.001) |

Figure 1 illustrates the effect modification of apoA-I on the relationship between impact of HDL-C and cardiovascular risk. At high concentrations of apoA-I (2 standard deviations above the mean), the relationship between HDL-C and risk is close to constant whereas at lower concentrations of apoA-I, cardiovascular risk increase with decreasing level of HDL-C.

**Figure 1.**
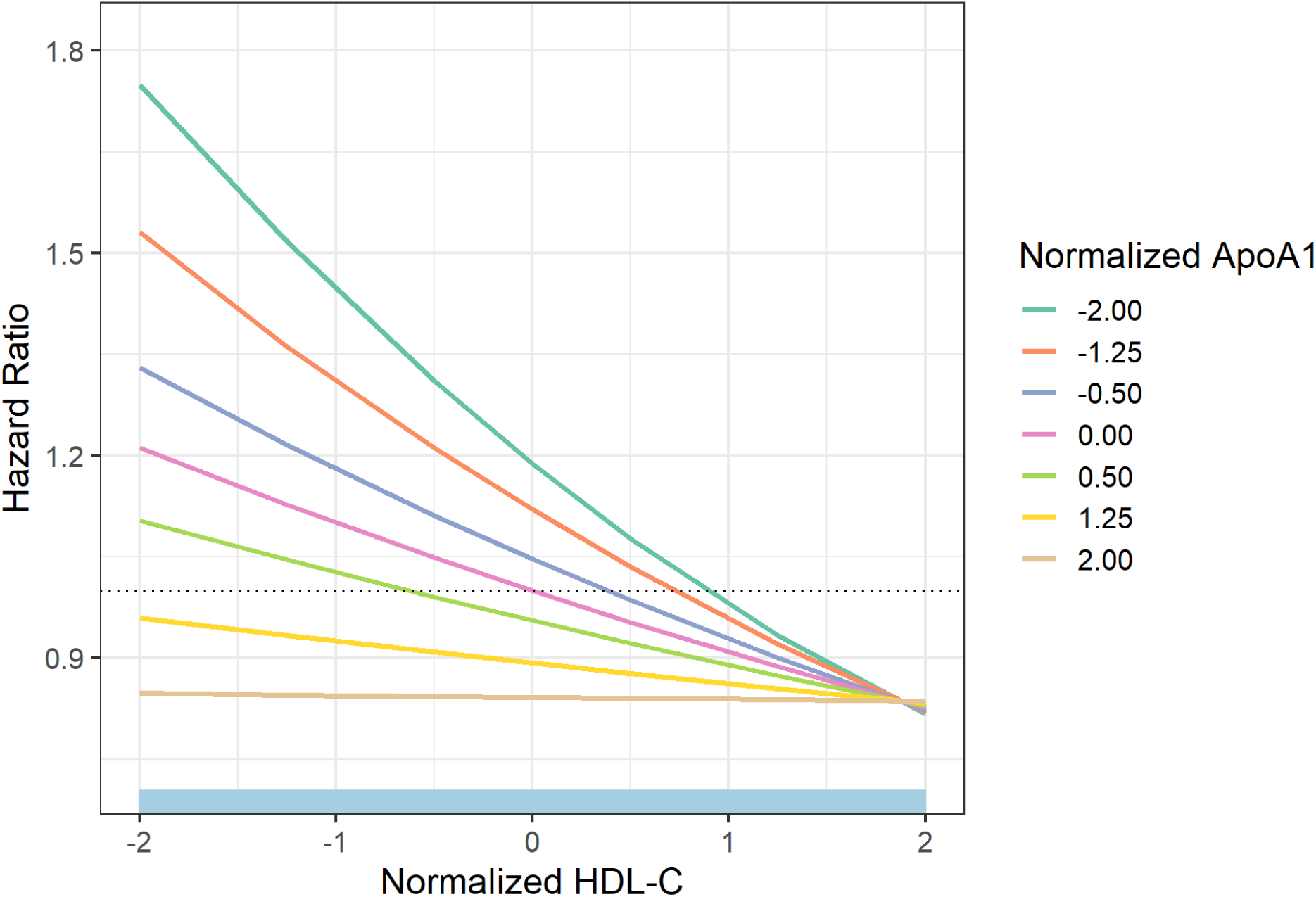
Interaction plot of normalized HDL-C and apoA-I versus cardiovascular risk. presents the results of the interaction of HDL-C and apoA-I. Both markers have been normalized to their respective per cent values. The hazard ratio (HR) of isopleths of apoA-I versus HDL-C are plotted.

### B. Discordance Analysis of HDL-C vs apoA-1 as Determinants of Cardiovascular Risk

In the Cox proportional hazards model with HDL-C and the apoA-I residual, HDL-C and the apoA-I residual were significantly associated with incident ASCVD (HR=0.85, 95% CI: 0.83-0.87, p<0.0001 and HR=0.96, 95% CI: 0.94-0.97, p<0.0001, respectively), as presented in Table 3. Similarly, in the Cox proportional hazards model with apoA-I and the HDL-C residual, apoA-I and the HDL-C residual were significantly associated with incident ASCVD (HR=0.85, 95% CI: 0.83-0.86, p<0.0001 and HR=0.98, 95% CI: 0.96-0.99, p<0.01, respectively).

**Table 3.** Hazard Ratios, 95% Confidence Intervals, and p-values for Incident Atherosclerotic Cardiovascular Disease in Cox Proportional Hazards Models in the Main Cohort.

| Variable | Hazard Ratio | 95% Hazard Ratio<br>Confidence Limits |  | P-value |
| --- | --- | --- | --- | --- |
| ApoA-1 | 0.85 | 0.83 | 0.86 | < 0.0001 |
| HDL-C Residual | 0.98 | 0.96 | 99 | 0.0098 |
| HDL-C | 0.85 | 0.93 | 89 | < 0.0001 |
| ApoA-1 Residual | 0.96 | 0.94 | 97 | < 0.0001 |
| ApoB | 1.14 | 1.12 | 1.16 | < 0.0001 |
| LDL-C<sup>1</sup> | 1.15 | 1.12 | 1.15 | < 0.0001 |
| Non-HDL-C<sup>1</sup> | 1.13 | 1.13 | 1.17 | < 0.0001 |
| InTG | 1.03 | 1.01 | 1.05 | 0.0107 |
| SBP | 1.19 | 1.17 | 1.20 | < 0.0001 |
| HbA1c | 1.08 | 1.07 | 1.09 | < 0.0001 |
| Age | 1.07 | 1.06 | 1.07 | < 0.0001 |
| Male | 1.70 | 1.65 | 1.75 | < 0.0001 |
| BMI | 1.08 | 1.07 | 1.10 | < 0.0001 |
| BP Medication | 1.52 | 1.46 | 1.57 | < 0.0001 |
| Diabetes Medication | 1.57 | 1.42 | 1.74 | < 0.0001 |
| Smoking | 1.24 | 1.20 | 1.28 | < 0.0001 |

To further investigate the effect on ASCVD of the relationship between apoA1 and HDL-C, we estimated the interaction between them as 1.05, 95%CI: (1.04, 1.06); p-value < 0.001.

### C. Relation of Plasma Triglycerides to HDL-C associated risk of cardiovascular disease

The fan-shaped relation of the concentration of apoA-I to the concentration of HDL-C is illustrated in Figure 2. As apoA-1 increases, so does HDL-C. But at any given concentration of apoA-1, there is variance in the concentration of HDL-C and this variance is related to the concentration of apoA-1, lower at lower levels of apoA-1 and higher at higher levels of apoA-1. The result is that the apoA-I and HDL-C plot fans out at higher concentrations of apoA-I. To investigate this relationship further, we postulated a decomposition of HDL-C into 3 components: apoA-I, (log) triglycerides, and residual of HDL-C regressed on apoA-I and log-triglycerides and estimated the relative value of these components in a multivariable-adjusted Cox regression model. The results are presented in Table 4.

**Figure 2.**
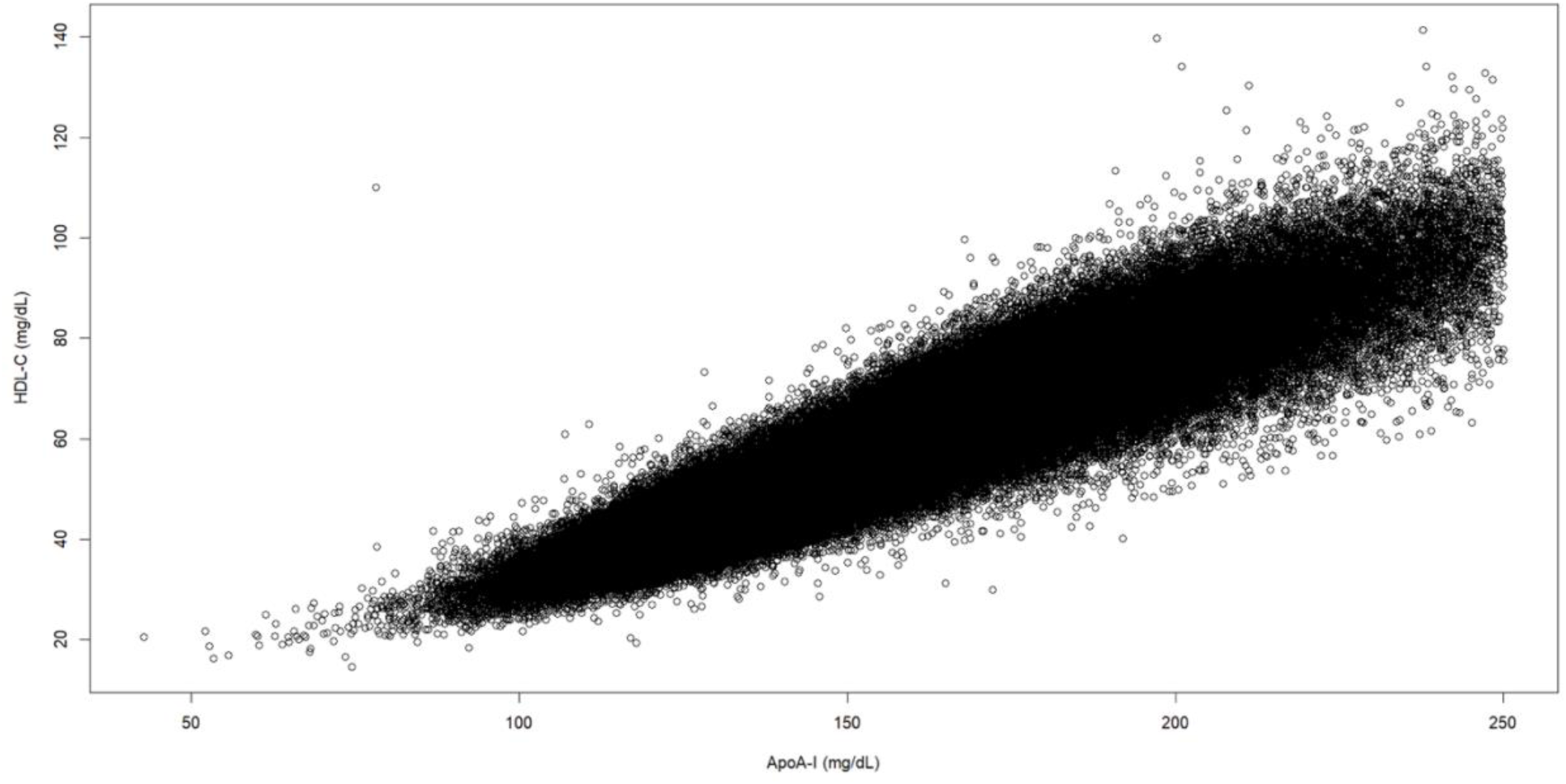
Plot of High-Density Lipoprotein-Cholesterol (HDL-C) Concentration (mg/dL) Against Apolipoprotein A-I (ApoA-I) Concentration (mg/dL) Plasma levels of apoA-I are plotted on the X-axis. Plasma levels of HDL-C are plotted on the Y axis. The appearance is fan-shaped with less variance in the level of HDL-C with lower levels of apoA-1 and greater variance of HDL-C with higher levels of apoA-I.

**Table 4.** Residual discordance analysis of HDL-C, apoA-I and ln TG on cardiovascular risk.

|  | HR (95%CI) | P | HR (95%CI) | P |
| --- | --- | --- | --- | --- |
| HDL-C |  |  | 0.836 (0.821 0.851) | < 0.001 |
| HDL-C residuals | 0.980 (0.965 0.995) | 0.010 |  |  |
| ApoA1 | 0.848 (0.833 0.863) | < 0.001 |  |  |
| ln TG | 1.041 (1.024 1.059) | < 0.001 |  |  |
| ApoB | 1.142 (1.125 1.16) | < 0.001 | 1.152 (1.136 1.169) | < 0.001 |
| SBP | 1.185 (1.168 1.202) | < 0.001 | 1.182 (1.165 1.199) | < 0.001 |
| HbA1c | 1.081 (1.07 1.092) | < 0.001 | 1.082 (1.071 1.093) | < 0.001 |
| Age | 1.067 (1.064 1.069) | < 0.001 | 1.067 (1.064 1.069) | < 0.001 |
| Male | 1.698 (1.645 1.752) | < 0.001 | 1.699 (1.646 1.754) | < 0.001 |
| BMI | 1.082 (1.066 1.098) | < 0.001 | 1.080 (1.064 1.097) | < 0.001 |
| BP med | 1.515 (1.463 1.568) | < 0.001 | 1.512 (1.46 1.565) | < 0.001 |
| DM med | 1.572 (1.416 1.744) | < 0.001 | 1.572 (1.417 1.744) | < 0.001 |
| Ever smoked | 1.239 (1.203 1.276) | < 0.001 | 1.238 (1.202 1.275) | < 0.001 |

When HDL-C alone is included in the multivariable model, its hazard ratio is 0.84 95% CI: (0.82, 0.85). Decomposing HDL-C into the 3 components, we observe that apoA-I accounts for the largest portion of the effect with a HR of 0.85 95%CI: (0.83, 0.86). Effects are much smaller for lnTG: 1.04 (1.02, 1.06) and residual of HDL-C: 0.98 95%CI: (0.96, 0.995).

## Discussion

This study has two principal findings. First, based on residual discordance analysis, HDL-C and apoA-I appear to have equally potent associations with cardiovascular risk: that is, the change in cardiovascular risk per standard deviation change of either marker in plasma is similar. However, notwithstanding that the concentrations of HDL-C and apoA-I are highly correlated and their overall HRs per SD change are similar, residual discordance analysis also demonstrates each enhances the association with risk of the other. Cardiovascular risk decreases as HDL-C concentration increases at the same concentration of apoA-I and, similarly, cardiovascular risk decreases as apoA-I concentration increases at the same concentration of HDL-C. Moreover, there was significant interaction between apoA-I and HDL-C and cardiovascular risk. The second principal finding is that decomposition of the relations of triglycerides, HDL-C and apoA-1 to risk demonstrated that triglycerides and residual HDL-C accounted for only a small portion of the potent inverse association of HDL-C to cardiovascular risk whereas apoA-I was the major determinant. This finding is relevant to the ongoing debate as to whether hypertriglyceridemia accounts for the cardiovascular risk that that has been attributed to HDL-C.

Overall, our findings are consistent with the relation of HDL-C to cardiovascular risk being the sum of two components. First, as apoA-1 increases, the range of values for HDL-C associated with a given value of apoA-1 increases. The result is that one component of the variance in HDL-C as apoA-I increases is due to a constant incremental increase in apoA-1-associated cholesterol. However, there is also a variable mass of cholesterol associated with apoA-I at any given concentration of apoA-I. This second factor accounts for the variance in concentration of apoA-I at any concentration of apoA-I and it is the combination of the two components that produces the total HDL-C in any individual. The significant interaction observed between HDL-C, apoA-1 and cardiovascular risk is consistent with both components contributing to the final predictive relation between HDL-C and risk with the predominant effect on risk being due the initial component.

However, in this study, as all others, there is an inverse relation between plasma triglycerides and HDL-C. With the negative results of Mendelian randomization analyses ^6–8^ and the failure of the HDL-C raising pharmacological trials, ^9^ support for a causal role for the triglycerides-rich particles increased, particularly with reports that such particles might be 4 to 5 times more atherogenic than LDL particles. ^12,13^ But if such studies are adjusted for the negative associations of risk with HDL-C, the positive associations with triglycerides are greatly attenuated.^14–16^

That there is an inverse relation between triglycerides and HDL-C is well-established and confirmed in the present study. Moreover, we have previously shown that HDL-C remains a significant predictor of risk after apoB and triglycerides are accounted for. ^14^ However, the relation of cardiovascular risk to apoA-I as opposed to HDL-C has not been studied in detail. Nor have the consequences of the HDL-C-lowering by triglycerides on the relation to cardiovascular risk been detailed. Accordingly, in this study, we partitioned the HDL-C attributable cardiovascular risk amongst triglycerides, apoA-I and HDL-C. Our analysis demonstrates that triglycerides and residual HDL-C account for only a minor portion of this risk with the majority being driven by apoA-I. Furthermore, in our study, the correlation coefficient between triglycerides and HDL-C-0.47-indicates less than 25% of the variance between the markers can be explained by the inverse relation. The correlation coefficient between triglycerides and apoA-1 is even lower-0.28. Thus, less than 10% of the variance in apoA-1 can be related to concurrent triglyceride levels. The effect of plasma triglycerides on the level of apoA-1 is, therefore, considerably less than the effect of plasma triglycerides on HDL-C. This indicates that other factors in addition to plasma triglycerides must affect the relation of HDL-C to apoA-I and the concentrations of both in plasma.

Why might the inverse relation to triglycerides be stronger for HDL-C than to apoA-I? We hypothesize this discordance arises because the decrease in cholesterol within HDL particles is the direct and immediate consequence of the replacement of a cholesterol ester molecule within the HDL particle by a triglyceride molecule mediated by cholesterol ester transfer protein. ^17^ Many, but not all, of these triglyceride molecules are subsequently hydrolyzed and the catabolism of apoA-I accelerated. ^17^ But the change in the concentration of HDL-C and the change in the concentration of concentration of apoA-I are two different processes that occur at two different rates and to two different extents. ^18^ And this, we believe, explains the different relationships between triglyceride and HDL-C versus triglycerides and apoA-I.

What then could account for risk relating both to apoA-I and the variance in HDL-C at any given level of apoA-I, with the former being more determinative of risk than the latter? The size, composition, and mass of cholesterol per HDL particle varies substantially. ^19^ The number of apoA-I molecules per HDL particles is variable, increasing from 2 to 4 as HDL particles become larger and cholesterol enriched. ^19,20^ Nevertheless, at any given total number of HDL particles, small and medium size HDL particles account for the great majority and their relative proportions do not vary substantially. ^18^ As an HDL particle acquires extracellular cholesterol, whether from a cellular or extracellular source, its mass of cholesterol will increase. ^21^ Similarly, the more HDL particles that can accept cholesterol from peripheral sources, the more cholesterol that can be taken up. ^22^ These basic metabolic features of HDL metabolism are consistent with HDL-C at any level of apoA-I being the sum of two components, the mass of cholesterol within the small and medium particles and a smaller and variable amount within the larger HDL particles. This two-component HDL-C model plus the evidence of significant interaction between HDL-C and apoA-I could account for our overall findings of strong relations of both apoA-I and HDL-C to cardiovascular risk as well as the smaller variance in risk due to variance in HDL-C at any given level of apoA-I. However, we acknowledge these are issues that require further study.

Finally, because our results are derived from an observational study, no causal inferences can be made. However, they do argue against assuming that HDL levels should not be taken into account when assessing the effects of triglycerides on cardiovascular risk. Moreover, given the consistency and potency of the inverse relation of HDL to cardiovascular risk, they support the continued search for a pathophysiological basis for this relation.

In summary, our discordance analysis demonstrates equivalent association with cardiovascular risk for apoA-I and HDL-C and that triglycerides account for only a minor portion of the HDL-associated risk of cardiovascular disease.

## Data Availability

Data will be provided in response to reasonable request.

## Abbreviations

HDL-C: high-density lipoprotein cholesterol
apoA-1: apolipoprotein A-1
apoB: apolipoprotein B
LDL-C: low density lipoprotein cholesterol
Non-HDL-C: non-high-density lipoprotein cholesterol

## Use of AI and AI-assisted Technologies Statement

AI technology was not used in the generation of this manuscript.

